# Maternal perception of child weight and concern about child overweight mediates the relationship between child weight and non-responsive feeding practices

**DOI:** 10.1101/2020.11.16.20232421

**Authors:** Jian Wang, Daqiao Zhu, Xuwen Cheng, Yicong LiuZhou, Bingqian Zhu, Scott Montgomery, Yang Cao

## Abstract

We aimed to examine the mediating effects of maternal perception of child weight (weight perception) and concern about overweight (weight concern) on the paths between child weight and non-responsive feeding practices. We recruited a convenience sample of 1164 mothers who were primary caregivers of preschool children. Child body mass index (BMI) Z-score was calculated to assess child weight. The Chinese version of the Child Feeding Questionnaire (C-CFQ) was used to measure four common non-responsive feeding practices, weight perception and weight concern. Structural equation modeling (SEM) was used to examine the associations between child BMI Z-scores, maternal feeding practices, and other covariates. Sixty percent of the mothers perceived their overweight/obese children as normal weight or even underweight. The disagreement between actual child weight and maternal weight perception was statistically significant (Kappa = 0.212, *P* < 0.001). SEM indicated that weight perception fully mediated the relationship between child BMI Z-scores and pressure to eat. Weight concern fully mediated the relationships between child BMI Z-scores and the other three feeding practices. The serial mediating effects of weight perception and concern were statistically significant for the paths between child BMI Z-score and monitoring (β = 0.035, *P* < 0.001), restriction (β = 0.022, *P* < 0.001), and food as a reward (β = -0.017, *P* < 0.05).

**Conclusion:** Child weight may influence maternal feeding practices through weight perception and concern. Thus, interventions are needed to increase the accuracy of weight perception, which may influence several maternal feeding practices and thereby contribute to child health.

**What is Known:**

- Non-responsive feeding practices may contribute to childhood obesity or eating disorders.
- Relationships between maternal weight perception and concern, child weight, and feeding practices have been mixed.

**What is New:**

- Child weight may influence maternal non-responsive feeding practices through maternal weight perception and concern.
- Interventions are needed to increase the accuracy of caregivers’ perception of child weight which may influence maternal feeding practices and thereby contribute to child health.

## 1. Introduction

Feeding practices refer to specific practices or strategies that parents employ to manage what, when and how much their children eat and shape their children’s eating patterns [1,2]. There are two types of parental feeding practices: non-responsive and responsive feeding [3-5]. Non-responsive feeding practices (also known as coercive control), such as applying pressure to eat, restricting food, monitoring consumption, and using food as a reward, is very common, especially in Chinese families [6-8]. These non-responsive feeding practices may contribute to the development of childhood obesity or long-term eating disorders [9-12].

Many studies have identified factors related to non-responsive feeding practices [13-17]. Among these factors, child weight has been an important factor [13,15-17]. Longitudinal studies indicated that mothers of children with higher body mass index (BMI) Z-scores were more likely to apply restrictive feeding practices [18-20] and monitor their children’s diet [18] and less likely to apply pressure to eat [18,20] than mothers of children with lower BMI Z-scores. However, it has been reported that parental perception of and concern about child weight (weight perception and weight concern) rather than actual child weight are associated with parental feeding practices [21-24]. For instance, Payne et al. [23] found that parents who were highly concerned about their child’s weight were more likely to use restrictive feeding than parents with lower levels of concern, while there was no significant association between child weight and feeding practices. Yilmaz et al. [24] found that parents were less likely to encourage their children to eat if they perceived them as overweight than if they perceived them as normal weight or underweight. In comparison, Freitas et al. [25] recently reported that maternal restrictive feeding practices were independently associated with mothers’ weight perception and concern as well as their child’s weight status. Overall, current evidence of the relationships between maternal weight perception and concern, actual child weight, and maternal non-responsive feeding practices has been inconsistent.

The inconsistent findings might be due to differences in factors included in the analyses (especially weight-related variables). Some studies did not examine maternal weight perception and/or weight concern and accordingly discovered only correlations between maternal feeding practices and child weight [15,26]. Other studies performed univariate analyses to investigate associations between factors of interest and parental feeding practices [15,24,27,28]. Collectively, there is a need to examine the intercorrelations between these variables.

Evidence-based theories of information processing and behaviorism learning, such as Gagne’s Information Processing Model [29], have suggested that cognitive changes (e.g., perception or concern) occur only when an objective stimulus (e.g., child weight) exists. The changes may then lead to specific behaviors (e.g., feeding practices). Similarly, Mareno proposed a middle-range explanatory theory in which the following elements occurred in order: parental beliefs and values about child body weight, parental perception of child weight, parental concern, and family lifestyle changes [30]. Therefore, caregivers’ weight perception and weight concern might mediate the association between child weight and feeding practices. A study of 7-to 9-year-old children in the U.K. reported that maternal concern about overweight fully mediated the relationship between child adiposity and the use of restrictive feeding practices [31]. In addition, previous research has indicated that child weight plays an important role in caregivers’ feeding practices [13,15-17,25].

Based on the above theories and empirical evidence, we proposed a hypothesized model (Figure 1). In this model, (1) child weight had a direct effect on four common non-responsive feeding practices, and (2) the associations between child weight status and these non-responsive feeding practices were mediated by maternal weight perception and weight concern. To explore the relationships proposed in the hypothesized model, we used structural equation modeling (SEM) while controlling for potential covariates, such as child age [14,16] and sex [14,16]. We also controlled for caregivers’ age [16,32] and weight classification based on BMI [16,17], education level [14,17], and family income [32,33].

**Figure 1.**
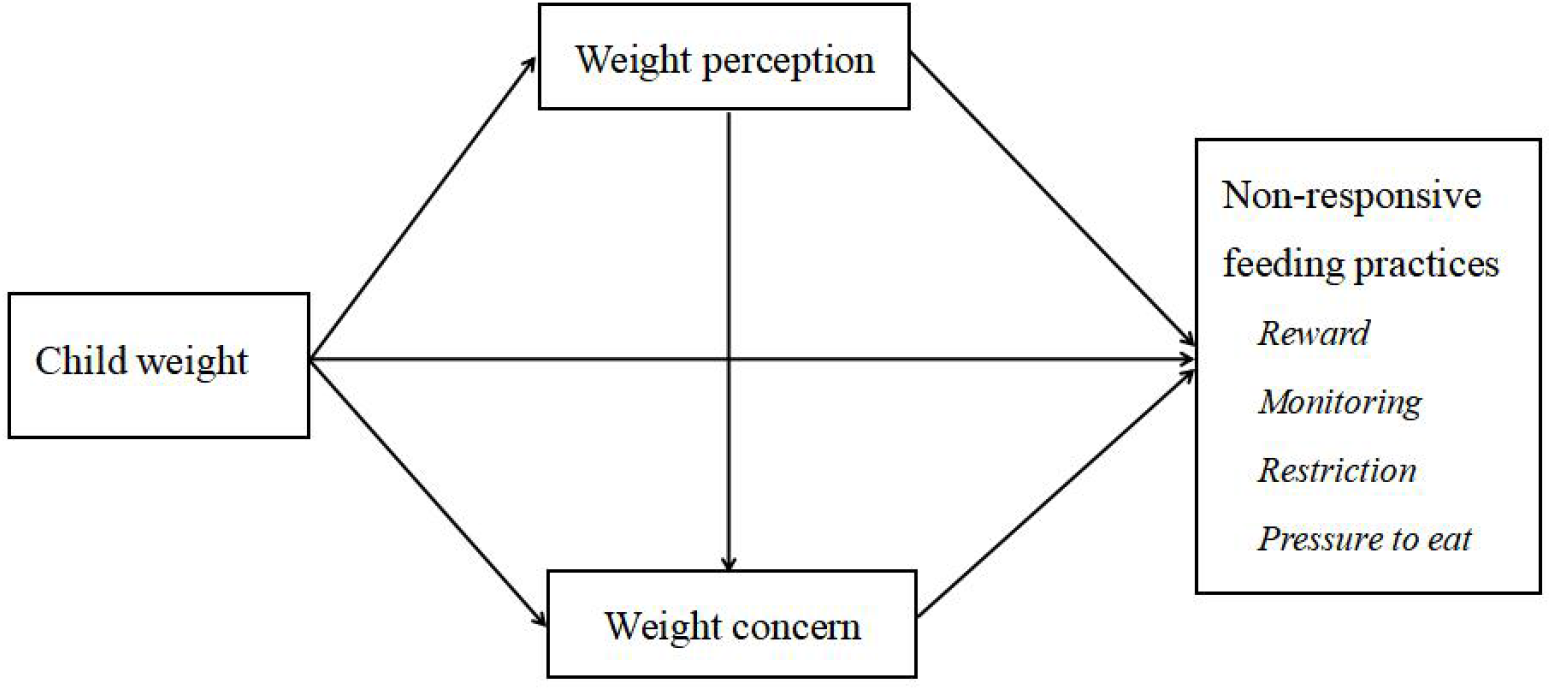
Hypothesized model for relationships between variables of interest. *Note*. Weight perception: perception of child weight; Weight concern: concern about child overweight.

## 2. Materials and Methods

### 2.1. Study design and participants

A correlational and cross-sectional design was used. This study was conducted between December 2017 and May 2020 in Shanghai, China. A convenience sample was selected from three public kindergartens located in three areas representing different economic levels in Pudong District, Shanghai. Only mothers were recruited because they are the primary caregivers of children typically in China. The required minimum sample size was 346, calculated using the formula below [34]:

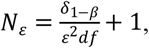

where *α* = 0.05; *β* = 0.90; *ε* = 0.05; *δ*_1–*β*_ ≈ 33.6; and *df* = 39.

A total of 1164 mothers were recruited. We excluded unqualified questionnaires, mothers who were not the primary caregivers, and children with diseases related to nutrition or extreme age-standardized BMI Z-scores. Finally, 1106 (95.02%) responses were included in this report.

Ethical approval was obtained from the Research Ethics Committee of Shanghai Jiao Tong University. Written informed consent was obtained from all participants.

### 2.2. Demographic and socioeconomic data

Demographic and socioeconomic data, including children’s age, sex and mothers’ age, weight, height, education level, and annual household income, were measured using a self-reported questionnaire. Children’s weight and height were measured and collected by trained health teachers in the kindergartens. Maternal weight status was classified as underweight (BMI < 18.5 kg/m^2^), normal weight (18.5kg/m^2^≤ BMI < 24.0 kg/m^2^), or overweight or obese (BMI ≥ 24.0 kg/m^2^) [35]. According to the World Health Organization (WHO) guidelines, child age-standardized BMI Z-scores were calculated using the software WHO Anthro (for 2-to 5-year-old children) and Young Growth Curve (for 5-to 6-year-old children). BMI Z-scores were categorized into three groups: underweight (Z-score < -2), normal weight (−2 ≤ Z-score≤ 1), and overweight or obese (Z-score > 1) [36].

### 2.3. Maternal feeding practices

Maternal feeding practices were evaluated using the Chinese version of the Child Feeding Questionnaire (C-CFQ). The C-CFQ assesses four types of non-responsive feeding practices: four items of monitoring (the extent to oversee their child’s eating [37]), six items of restriction (strict limitations on the child’s access to foods or opportunities to consume unhealthy foods [38]), four items of pressure to eat (insists, demands, or physically struggles with the child in order to get the child to eat more food [37-39]), and two items of food as a reward (use of desired foods as a way to regulate the child’s eating or behaviors [39]). Each item was rated on a 5-point Likert scale. The response options for each item were “always,” “usually,” “sometimes”, “almost never”, and “never”. Each subscale was calculated by averaging the scores of all the items in that subscale. The C-CFQ demonstrated good internal consistency reliability in the current study (Cronbach’s α = 0.748-0.890). The questionnaire also showed good construct validity [40,41].

### 2.4. Maternal perception of child weight (weight perception) and concern about overweight (weight concern)

Weight perception was assessed by asking “How would you describe your child’s weight?”. The responses included “very underweight,” “slightly underweight,” “normal weight,” “slightly overweight,” and “very overweight.” Weight concern was assessed by asking “How concerned are you about your child becoming or staying overweight in the future”. The responses included “unconcerned,” “slightly concerned,” “concerned,” “fairly concerned,” and “very concerned.” Both items have been used and validated in previous studies [22,31].

### 2.5. Statistical analysis

SPSS Statistics 24.0 and SPSS Amos 24.0 for Windows (IBM Corp, Armonk, NY, USA) were used for data coding, cleaning, and analysis. Descriptive statistics were used to describe the participants’ characteristics. The agreement between maternal weight perception and reported child weight was assessed using the Kappa statistic. Pearson’s correlation analysis was used to explore the relationships between continuous variables. All the studied variables were approximately normally distributed with skewness and kurtosis below 2.0. Parameters for the hypothetical path model were estimated using structural equation modeling (SEM) via maximum likelihood estimation. The path analysis was conducted, and goodness of fit of the models was evaluated using the following indices and cut-offs: χ^2^ between 1.0 and 2.0, goodness of fit index (GFI) and comparative fit index (CFI) > 0.90, root mean square error of approximation (RMSEA) < 0.06, root mean square residual (RMR) < 0.08, and a small Akaike information criterion (AIC) [40,41]. In the final model, we controlled for demographics that were significantly associated with maternal feeding practices (*P* < 0.05). The following conditions suggest a significant mediating effect: (1) the initial predictor (child BMI Z-score) was associated with the outcome (feeding practices) and the proposed mediators (perception or concern); (2) the mediators were associated with the outcome; and (3) the effect of the predictor on the outcome decreased substantially once the mediator was added as a second predictor in the model. To control for type I errors, the bootstrap method was used when examining the mediating effect of weight perception and weight concern [42]. Statistical significance was set at *P* < 0.05 (two-sided).

## 3. Results

### 3.1. Demographic and socioeconomic characteristics

The demographic characteristics of the participants are shown in Table 1. Two hundred nineteen (19.8%) preschool children were reported to be overweight or obese. A few demographic variables were statistically significantly related to mothers’ feeding practices (not shown). Maternal age was negatively associated with the use of food as a reward (r = -0.099, *P* = 0.001). Maternal educational level was negatively associated with applying pressure to eat (r = -0.121, *P* = 0.002) and the use of food as a reward (r = -0.087, *P* = 0.025). Child age was negatively associated with maternal monitoring of the child’s diet (r = -0.111, *P* < 0.001), applying pressure to eat (r = -0.067, *P* = 0.025) and restriction of food (r = -0.060, *P* = 0.045). Annual household income showed a positive relationship with maternal monitoring of the child’s diet (r = 0.084, *P* = 0.005). These variables were added to the model as covariates.

**Table 1.** Demographic characteristics of the participants (*n* = 1106)

| | $n$ (%) or (mean ± SD) |
| --- | --- |
| **Child characteristics** |  |
| Age | 4.56 ± 1.35 |
| Sex |  |
| Boys | 588 (53.3) |
| Girls | 516 (46.7) |
| Weight status |  |
| Underweight: BMI Z-score < -2 | 44 (4.0) |
| Normal weight: $-2 \leq$ BMI Z-score $\leq 1$ | 843 (76.2) |
| Overweight or Obesity: BMI Z-score > 1 | 219 (19.8) |
| **Mother characteristics** |  |
| Age | 34.66 ± 3.96 |
| Weight status |  |
| Underweight: BMI < 18.5 kg/m<sup>2</sup> | 111 (10.2) |
| Normal weight: 18.5 kg/m<sup>2</sup> ≤ BMI < 24.0 kg/m<sup>2</sup> | 784 (71.8) |
| Overweight or Obesity: BMI ≥ 24.0 kg/m<sup>2</sup> | 197 (18.0) |
| Education level |  |
| College or higher | 872 (78.8) |
| Senior high school or below | 234 (21.2) |
| Household income/year |  |
| Above average (> CNY 300,000) | 638 (57.7) |
| Below average (≤ CNY 300,000) | 468 (42.3) |

Notes. SD: standard deviation; BMI: body mass index; BMI Z-score: body mass index Z-score; CNY: Chinese Yuan

### 3.2. Agreement between maternal perception of child weight (weight perception) and actual child weight status

Since few mothers perceived their children’s weight as very overweight or very underweight, these two statuses were combined with overweight and underweight, respectively. Three categories (perceived underweight, normal weight, and overweight) were used in the final analysis. 14.1% of mothers felt that their children weighed too much, 59.0% reported that their children weighted a normal weight, and 26.9% felt that their children were underweight. The agreement between maternal weight perception and actual child weight status was weak (*Kappa* = 0.212, *P* < 0.001) (Table 2). A total of 660 (59.67%) mothers properly perceived their children’s weight status, and 378 (34.18%) mothers underestimated their children’s weight status. Most mothers of overweight/obese children felt their children were normal weight (*n* = 106, 48.4%) or even underweight (*n* = 9, 4.1%) (Table 2).

**Table 2.** Agreement of actual child weight status with maternal weight perception

| Actual child weight status<br>(BMI Z-score) | Maternal weight perception ($n = 1106$) | | | |
| --- | --- | --- | --- | --- |
|  | Underweight | Normal weight | Overweight/Obesity | Total |
| Underweight | 26 (59.1%) | 16 (36.4%) | 2 (4.5%) | 44 (3.98%) |
| Normal weight | 263 (31.2%) | 530 (62.9%) | 50 (5.9%) | 843 (76.22%) |
| Overweight/Obesity | 9 (4.1%) | 106 (48.4%) | 104 (47.5%) | 219 (19.80%) |
| Total | 298 (26.9%) | 652 (59.0%) | 156 (14.1%) | 1106 |

*Notes.* BMI Z-score: body mass index Z-score; Weight perception: maternal perception of child weight; $Kappa = 0.212$ , $P < 0.001$ .

### 3.3. Description of variables included in the path analysis

Of the mothers, 74.1% were not concerned about their child becoming overweight, 18.4% reported some concern, and 7.5% reported great concern. The descriptions of weight perception, concern, and feeding practices are shown in Table 3. The mean scores of weight perception (2.84) and concern (1.85) were both lower than the median level of 3. Pressure to eat and food as a reward were approximately at the median level. Slightly higher mean scores were reported for restriction and monitoring.

**Table 3.** Description of weight perception, concern and feeding practices (*n* = 1106)

| Variables | Range | Item numbers | Mean | SD |
| --- | --- | --- | --- | --- |
| Weight perception | 1-5 | 1 | 2.84 | 0.70 |
| Weight concern | 1-5 | 1 | 1.85 | 1.03 |
| Pressure to eat | 1-5 | 4 | 2.97 | 0.79 |
| Food as a reward | 1-5 | 2 | 3.38 | 0.76 |
| Restriction | 1-5 | 6 | 3.88 | 0.61 |
| Monitoring | 1-5 | 4 | 3.97 | 0.82 |

*Notes.* Weight perception: maternal perception of child weight; Weight concern: maternal concern about child overweight.

**Table 4.** Correlations of the key variables (*n* = 1106)

|  | Monitoring | Pressure to eat | Food as a reward | Restriction | BMI Z-score | Weight perception |
| --- | --- | --- | --- | --- | --- | --- |
| Pressure to eat | 0.142** |  |  |  |  |  |
| Food as a reward | -0.097** | 0.144** |  |  |  |  |
| Restriction | 0.445** | 0.254** | 0.026 |  |  |  |
| BMI Z-score | 0.042 | -0.084** | -0.025 | 0.052 |  |  |
| Weight perception | 0.032 | -0.098** | -0.038 | 0.065* | 0.514** |  |
| Weight concern | 0.124** | -0.058 | -0.065* | 0.108** | 0.390** | 0.528** |

*Notes:* BMI Z-score: body mass index Z-score; Weight perception: maternal perception of child weight; Weight concern: maternal concern about child overweight. \* $P < 0.05$ , \*\* $P < 0.01$ .

### 3.4. Relationships between variables in the path analysis

The bivariate associations between the four feeding practices (monitoring, pressure to eat, use food as a reward, and restriction), BMI Z-score, and weight perception and concern are shown in Table Most of the associations were statistically significant.

### 3.5. Path analysis

Based on the hypothesized model (Figure 1) and the above bivariate correlation analyses, the following variables were included in the initial model: BMI Z-score, weight perception and concern, monitoring, restriction, pressure to eat, food as a reward, child age, maternal age, maternal education level, and annual family income (Figure 2). The structural model revealed a good fit of the data: *χ*^2^_(32)_=86.297 (*P* < 0.001), GFI = 0.987, CFI = 0.961, RMSEA = 0.046, RMR = 0.035, and AIC = 176.297. In the initial model, three direct paths were not significant. We removed the three non-significant paths respectively to obtain the most parsimonious model. After simplifying the model, all the remaining statistically significant paths were entered into a final model to assess the relative importance of the direct and indirect effects of BMI Z-score on maternal feeding practices.

**Figure 2.**
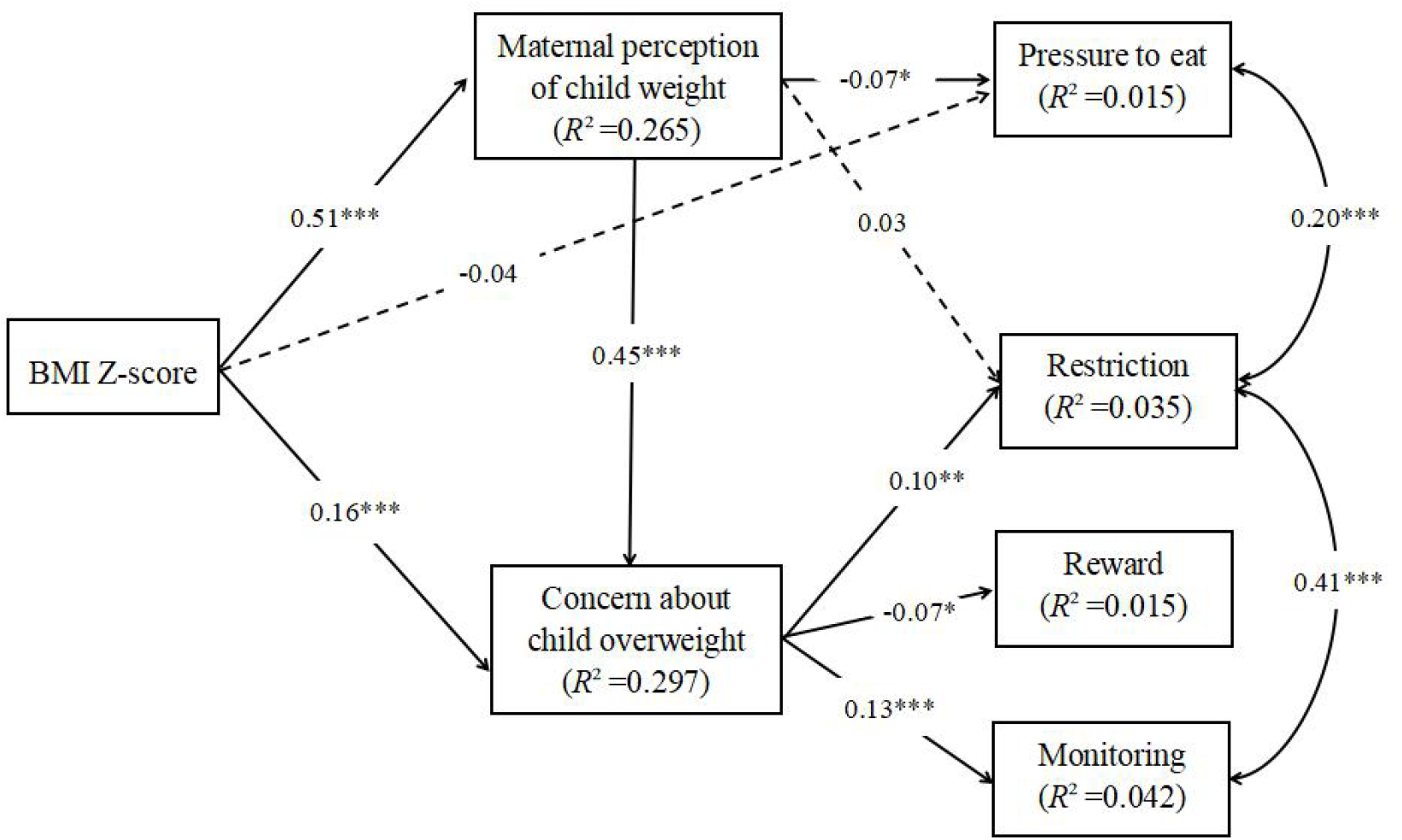
Initial model of the relationships between the study variables. *Notes*: Paths were adjusted for child age, maternal age, maternal education level, and annual household income. Arrows represent directions. Significant paths are shown as solid lines, and non-significant paths are shown as dashed lines. BMI Z-score: body mass index Z-score; Weight perception: maternal perception of child weight; Weight concern: maternal concern about child overweight. ^***^*P* < 0.001; ^**^*P* < 0.01; ^*^*P* < 0.05.

The final model is illustrated in Figure 3. It showed a better fit than the initial model, with the following parameters: *χ*^2^_(34)_ =89.109 (*P* < 0.001), GFI = 0.986, CFI = 0.960, RMSEA = 0.038, and RMR = 0.046. The AIC value was lower in the current model (153.109) than in the previous model. The model accounted for 1.5%, 4.1%, 3.4%, and 1.5% of the variance in food as a reward, monitoring, restriction, and pressure to eat, respectively. All the direct paths in the final model were statistically significant. Specifically, weight concern was associated with monitoring (standardized *β* = 0.13, *P* < 0.001), restriction (standardized *β* = 0.12, *P* < 0.001), and food as a reward (standardized *β* = -0.07, *P* < 0.05). The standardized *β* of the direct path from weight perception to pressure to eat was -0.10 (*P* < 0.001).

**Figure 3.**
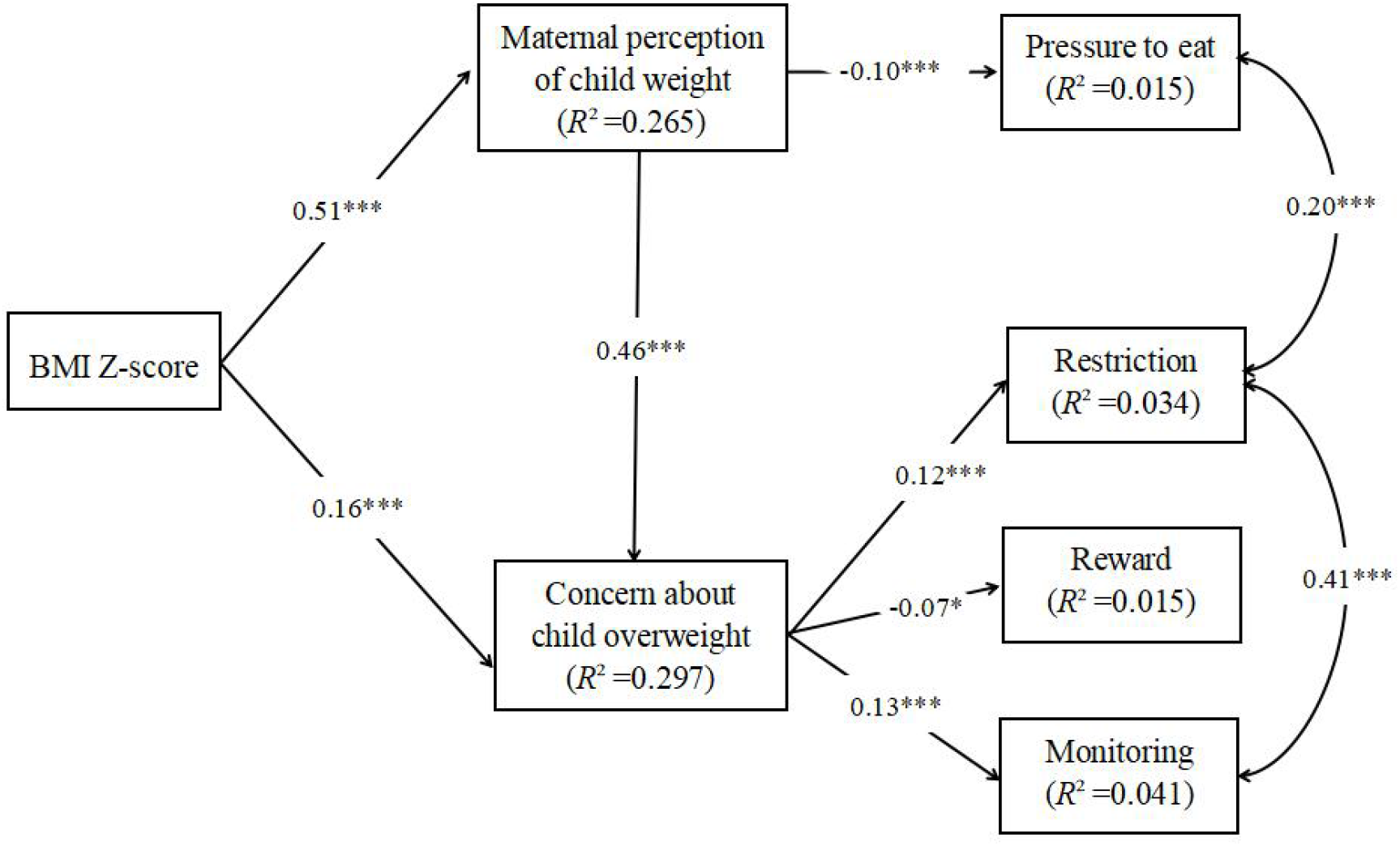
Final model of the relationships between the study variables. *Notes*: BMI Z-score: body mass index Z-score; Weight perception: maternal perception of child weight; Weight concern: maternal concern about child overweight. ^***^*P* < 0.001; ^**^*P* < 0.01; ^*^*P* < 0.05.

### 3.6. Indirect effects of child weight and maternal perception of child weight (weight perception) on maternal feeding practices

The indirect effects of child weight and weight perception on maternal feeding practices are presented in Table 5. When all the variables were included in the model, both paths through a single mediation (indirect effects 1-4 in Table 5) and a serial mediation (indirect effects 5-7 in Table 5) were statistically significant. The total indirect impact on maternal feeding practices was also statistically significant (*P* < 0.05). The mediation of weight concern (i.e., M2 in Table 5) between weight perception (i.e., M1 in Table 5) and food as a reward, restriction, and monitoring (i.e., Y1, Y2, and Y3 in Table 5) was also statistically significant.

**Table 5.** Indirect effects of child weight and weight perception on maternal feeding practices

| Effect | $\beta$ | Boot SE | Bootstrapping 95%<br>Confidence Interval | | Z |
| --- | --- | --- | --- | --- | --- |
|  |  |  | Lower | Upper |  |
| Indirect 1: $X \rightarrow M1 \rightarrow Y4$ | -0.034 | 0.010 | -0.055 | -0.014 | 3.4** |
| Indirect 2: $M1 \rightarrow M2 \rightarrow Y1$ | -0.033 | 0.016 | -0.066 | -0.002 | 2.06* |
| Indirect 3: $M1 \rightarrow M2 \rightarrow Y2$ | 0.044 | 0.011 | 0.022 | 0.067 | 4*** |
| Indirect 4: $M1 \rightarrow M2 \rightarrow Y3$ | 0.069 | 0.015 | 0.040 | 0.100 | 4.6*** |
| Indirect 5: $X \rightarrow M1 \rightarrow M2 \rightarrow Y1$ | -0.017 | 0.008 | -0.034 | -0.001 | 2.125* |
| Indirect 6: $X \rightarrow M1 \rightarrow M2 \rightarrow Y2$ | 0.022 | 0.006 | 0.011 | 0.034 | 3.67*** |
| Indirect 7: $X \rightarrow M1 \rightarrow M2 \rightarrow Y3$ | 0.035 | 0.008 | 0.020 | 0.052 | 4.375*** |

Notes: N = 1106. Number of bootstrap samples for bias-corrected bootstrap confidence intervals: 10000. X = BMI Z-score, M1 = Weight perception, M2 = Weight concern, Y1 = Reward, Y2 = Restriction, Y3 = Monitoring, Y4 = Pressure to eat; SE, standard error; \* $P < 0.05$ , \*\* $P < 0.01$ , \*\*\* $P < 0.001$ .

## 4. Discussion

In this study, we examined the mediating roles of maternal perception of child weight and concern about overweight on the association between child weight and non-responsive feeding practices. The results from the SEM largely supported our hypotheses.

In this study, although the child BMI Z-score was significantly related to pressure to eat in the bivariate analysis, it was not a direct predictor in the SEM. Based on the bootstrapping results, child BMI Z-score influenced mothers’ feeding practices via three paths: two full single mediating effects of weight perception and concern and a serial multiple mediating effects of weight perception and concern. Specifically, the relationship between child BMI Z-score and pressure to eat was fully mediated by weight perception. This finding suggested that mothers of children with higher BMI Z-scores were more likely than those of children with lower BMI Z-scores to perceive their children as having a high weight, which in turn decreased the likelihood of them forcing their children to eat. Additionally, weight concern fully mediated the associations between child BMI Z-score and maternal restriction, monitoring, and use of food as a reward. In this case, mothers whose children were overweight or obese were more likely than those whose children were normal or underweight to report concerns and monitor or restrict their children’s intake rather than reward them for eating. In addition, mothers of children with higher BMI Z-scores were more likely to perceive their children as having a high weight status and express concern about their children being overweight or obese, which ultimately affected their feeding practices, such as restricting the consumption of unhealthy food.

Previous studies have assessed the correlations between caregivers’ feeding practices and weight-related variables [13,21,24,27]; however, few have explored their interrelationships. A population-based cohort study showed that mothers’ concerns about their children’s weight mediated the relationship between child BMI Z-score at 4 years and restrictive feeding at 10 years [43]. Our findings are consistent with previous evidence [21,22,31] and in accordance with Costanzo and Woody’s suggestion [44] that parents should use controlled feeding practices when they are concerned about their child’s weight. Therefore, although the results of this study supported our hypotheses, the mediating effect of weight perception and concern warrants further investigation.

In the current study, maternal perception of child weight was a mediator of child BMI and maternal feeding practices. This finding emphasizes the importance of accurate perception of child weight. Only when mothers recognize their preschool children’s weight problems do they implement effective feeding practices to control their children’s weight. In this study, we found a high percentage of misperception of child weight, with mothers tending to underestimate their children’s weight. The results are consistent with recent findings [45,46]. The inability to accurately identify child overweight/obesity is problematic given that substantial evidence suggests that weight perception is critical in motivating health-related behavioral changes [47-49]. Thus, mothers who consider their overweight/obese children as normal weight or even underweight might not be concerned about their children being overweight or obese and therefore adopt inappropriate feeding practices, such as forcing children to eat more. This practice may further influence a child’s development and growth, especially their weight.

Although our model showed a good fit of the data, the variance in feeding practices accounted for by the predictors was relatively low. The low variance was in line with other studies [21,31]. For instance, Gregory et al. [21] found that maternal concern about child overweight was a significant predictor, explaining an additional 2% of the variance in restriction, while food responsiveness explained 7%. It is likely that weight perception and concern were not major contributing factors of maternal feeding practices. Another possible explanation is that important factors related to maternal feeding practices might have been missed [50]. Compared to mothers recruited in other studies [17,25,31], mothers in our study showed a relatively low level of weight concern. Additionally, the correlations between weight concern and maternal feeding practices were weak. These findings indicate that mothers in our study might have used specific feeding practices to address their concerns about their children’s undereating habits and underweight [13,21,51]. For instance, it has been indicated that maternal use of pressure to eat is independently associated with maternal concern about their child eating too little [13,51]. Similarly, Gregory et al. [21] found that maternal concern about child underweight was the strongest independent predictor of pressure to eat, contributing to 15% of the variance. In contrast, mothers tend to consider a relatively high child weight as an indicator of good health, especially in China [52-54]. They thus may be less concerned about child overweight than mothers in other countries. Moreover, monitoring or restricting unhealthy food intake may be associated with maternal concern about pediatric health problems, resulting in a decreased/lack of appetite or chronic diseases, such as obesity [49,55,56].

To the best of our knowledge, this study was among the first to examine the structural relationships between child weight, mothers’ weight perception and weight concern, and non-responsive feeding practices. The significant paths and well-fitted model provided further empirical support for the hypothetical model. However, this study has limited generalizability because of the sampling method used (e.g., a convenience sampling of only mothers). Although the prevalence of overweight or obese children in our study was similar to that in a large-scale study conducted in Shanghai, China [57], random samples from multiple sites are needed in the future. Another limitation was the correlational design, which precluded us from making causal inferences. Additionally, self-reported information may be subject to recall bias. Finally, we focused on only non-responsive feeding practices, whereas other positive approaches of feeding (e.g., rules, limits, and modeling) were not assessed. Thus, longitudinal studies using comprehensive questionnaires are needed to confirm findings from this study.

## 5. Conclusion

In conclusion, maternal perception of child weight and concern about overweight rather than actual child weight may be related to mothers’ feeding practices. Mothers’ perception of their children’s weight not only influenced their non-responsive feeding practices directly but also had an indirect effect via concern about child overweight. Thus, non-responsive feeding practices associated with childhood obesity should first address maternal perception of child weight. Managing concern about child overweight may be a potential intervention. Future studies should focus on correcting caregivers’ misperception about child weight. Interventions designed to improve caregivers’ accurate assessments of child weight are needed so that they can adopt appropriate feeding practices, thereby optimizing child health.

## Data Availability

The data that support the findings of this study are available from the corresponding author, upon reasonable request.

## Acknowledgements

We would like to thank all of the child healthcare workers at Jinyang Community Health Service Center for their help with data collection.

## Author contributions

Data collection: X.C., Y.C., L.Z., and D.Z.; Formal analysis: J.W.; Investigation: X.C., Y.C., L.Z., and D.Z.; Methodology: D.Z., and Y.C.; Project administration: D.Z.; Resources: D.Z.; Software: J.W.; Supervision: D.Z., S.M. and Y.C.; Visualization: J.W.; Writing – original draft: D. Z, J.W., and Y.C.; and Writing – review and editing: J.W., X.C., B.Z., D.Z., S.M., and Y.C.

## Compliance with ethical standards

### Ethical approval

Ethical approval was obtained from the Research Ethics Committee of Shanghai Jiao Tong University. Written informed consent was obtained from the mothers.

### Consent for publication

All caregivers understood and agreed that their information could be published in any journal. The authors declare that there is no conflict of interest regarding the publication of this article.

## Abbreviations

AIC: Akaike information criterion
BMI: Body mass index
BMI Z-score: body mass index Z-score
C-CFQ: Chinese version of the Child Feeding Questionnaire
CFI: comparative fit index
CNY: Chinese Yuan
GFI: goodness of fit index
RMR: root mean square residual
RMSEA: root mean square error of approximation
SD: standard deviation
SE: standard error
SEM: structural equation modeling
Weight concern: maternal concern about child overweight
Weight perception: maternal perception of child weight

